# Machine Learning Phenotyping of Autonomic Stress from Daily Temperature and Ecological Assessments

**DOI:** 10.1101/2025.11.11.25339837

**Authors:** Alvaro Alaor da Silva

## Abstract

**Background:** Artificial intelligence applications for preventive stress monitoring remain limited by dependence on expensive continuous biosensors. We developed and validated an AI-based framework for automated phenotyping of stress-energy responses from accessible smartphone-based circadian temperature monitoring and cognitive-autonomic assessments, enabling scalable population health monitoring without wearable devices.

**Methods:** This 15-day prospective observational study collected 239 daily observations from 16 adults (age 58.35±7.8 years; 100% adherence). Daily axillary temperature oscillation (ΔT = night-minus-morning), a 6-item cognitive-autonomic index (MiSBIE Brief-6), morning light exposure, and screen time were analyzed using unsupervised K-means clustering. A Composite Stress Load (CSL) index integrating subjective stress (40%), thermal variance (30%), and pain (30%) was computed. Cluster validation employed silhouette analysis, Gap statistics, and Hopkins test.

**Results:** Unsupervised machine learning identified three distinct stress-energy phenotypes (k=3; silhouette=0.75; Gap p<0.001): Cluster 1 (Low ΔT/High Recovery; n=87; ΔT=-0.19±0.09°C; MiSBIE-delta=+1.84±0.62), Cluster 2 (Neutral/Intermediate; n=98; ΔT=+0.00±0.07°C; MiSBIE-delta=+1.12±0.51), and Cluster 3 (High ΔT/Minimal Recovery; n=54; ΔT=+0.21±0.10°C; MiSBIE-delta=+0.41±0.68). Elevated ΔT strongly correlated with CSL (r=0.52; p<0.001). AI-derived phenotypes predicted 78% of thermal stability variance (R^2^=0.78; p<0.001). Morning light >15 minutes reduced ΔT (β=-0.24°C; p=0.002).

**Conclusions:** This validated AI framework achieves automated stress phenotyping at <$5 per participant versus $200-500 for wearables, supporting early identification of elevated allostatic load aligned with the Energy Resistance Principle. Longitudinal phenotype tracking enables predictive early warning and individualized exercise optimization in real-world settings, advancing health equity in preventive monitoring for resource-limited contexts. Integration into public health systems serving millions (e.g., Brazil’s SUS) could enable anticipatory care delivery, improving quality of life through early intervention before clinical deterioration

**Highlights:**

- K-means clustering identified 3 autonomic phenotypes (silhouette=0.75)
- ΔT>0°C predicts stress load elevation (r=0.52, p<0.001)
- MiSBIE-6 explains 78% of thermal variance (R^2^=0.78, p<0.001)
- Morning light reduces ΔT by 0.24°C; screens increase it 0.19°C/hour
- Smartphone-based framework enables scalable stress phenotyping[AA1]

## Introduction

Artificial intelligence (AI) is transforming preventive healthcare by enabling automated phenotyping and early detection of health risks from accessible data (33–35). Machine learning (ML) approaches, particularly unsupervised clustering, can identify disease subtypes and risk profiles without requiring expensive diagnostic infrastructure or manual labeling (34). However, AI applications in stress and autonomic monitoring remain limited by dependence on costly continuous biosensors—heart rate variability (HRV) monitors cost $200-500 USD and require specialized wearables with limited accessibility in resource-constrained settings (36). This technological barrier restricts population-scale stress monitoring to high-resource contexts, perpetuating health inequities.

This study presents an AI-based framework for stress-energy phenotyping using machine learning analysis of daily temperature oscillations (ΔT) combined with ecological momentary assessment of cognitive-autonomic states. By replacing expensive continuous monitoring with accessible daily measurements, this approach enables scalable AI-driven stress phenotyping at <$5 per participant, democratizing preventive health monitoring across diverse socioeconomic contexts.

Daily axillary temperature follows a circadian pattern governed by the suprachiasmatic nucleus (SCN) and peripheral tissue clocks expressing canonical clock genes (14). Morning light exposure advances circadian phase and stabilizes autonomic rhythms (3), whereas insufficient morning light or prolonged evening screen use promotes circadian misalignment, melatonin suppression, and reduced nocturnal vagal recovery (4,14,19).

The energetic model of allostatic load (EMAL), originally proposed by McEwen & Stellar and extended by Goldstein, delineates the point at which energy allocated to stress responses competes with growth, maintenance, and repair processes. When chronic, this energetic shift associates with compensatory hypermetabolism, cumulative physiological wear, and increased vulnerability to stress-related disease (15,16). The Energy Resistance Principle (ERP) extends EMAL into mitochondrial physiology, proposing that ‘energy resistance’ (éR) emerges when the electron transport system becomes less efficient under sustained allostatic load (10,21).

Under high éR, the same metabolic substrate yields less usable ATP per unit of oxygen consumed, necessitating increased metabolic effort—a state that, if the ERP framework holds, should manifest as impaired nocturnal heat dissipation. When daily energetic demand exceeds the organism’s capacity to dissipate thermal and metabolic entropy during the recovery window, ΔT remains elevated (>0°C), signaling incomplete return to baseline and accumulation of allostatic debt.

Picard et al. demonstrated that individuals with higher prior allostatic load exhibited 72-hour slower recovery and +45% GDF15 elevation following identical training stimuli (10). This finding illustrates that the capacity to tolerate a given stressor depends not on stressor intensity per se, but on the pre-existing energetic budget available for dissipation—analogous to the “recovery capacity” indexed by ΔT. When this capacity is exceeded, the organism enters a desadaptive state characterized by elevated éR, entropic accumulation, and progressively compromised cellular regeneration.

The Mitochondrial Stress, Brain Imaging, and Epigenetics (MiSBIE) program demonstrates individuals vary markedly in their psychobiological stress-reactivity profiles, mitochondrial stress sensitivity, and capacity for rhythmic recovery (11,12). The MiSBIE framework—a six-domain cognitive–autonomic composite including mental clarity, vitality, emotional balance, tension release, interoception, and subjective rhythmic synchrony—has shown strong internal consistency and convergence with autonomic and inflammatory markers in previous work (11).

Despite robust laboratory evidence, a critical gap exists: ecologically valid, low-cost metrics capable of monitoring éR, allostatic load, and autonomic coherence in real-world settings. Recent evidence positions daily temperature difference (ΔT) as an integrative thermodynamic component of nocturnal vagal recovery, serving as an indirect HRV marker in non-instrumented settings (17,18). Hasselberg et al. showed nocturnal ΔT strongly correlated with HRV indices in free-living adults (17); Blatteis reported ΔT reflects vagally induced heat loss; Romeijn & Van Someren found strong negative correlation between skin temperature gradient and HRV power (r=−0.76; p<0.001) (18). These findings establish ΔT as a thermodynamic proxy for vagal recovery with ecological validity superior to wearable HRV in unconstrained settings (17,18).

Our central hypothesis is that ΔT ≤ 0 °C reflects low éR and efficient allostatic load dissipation via nocturnal vagal repair; whereas ΔT > 0 °C reflects high éR, hypermetabolism, autonomic instability, and cellular energetic strain. This thermal metric, combined with MiSBIE Brief-6 and a composite stress load index (CSL), may identify the allostasis-to-overload transition—the point where identical stressors become maladaptive.

## METHODS

### Study Design

Prospective observational longitudinal study (15 days) with 100% adherence, conducted according to STROBE standards. The design incorporated ecological assessment of natural daily stressors and their influence on circadian body temperature, consistent with known SCN and clock-gene regulation of thermal rhythmicity. The study included MiSBIE multi-level assessment in alignment with its established psychobiological framework.

### Population and Sample

Sixteen healthy adults participated (9 women, 7 men; mean age 58.35 ± 7.8 years). Exclusion criteria included circadian rhythm disorders (MEQ <31 or >69), use of chronotropic medications, or recent changes in medication. Participants were recruited through professional networks. Completion rate was 100%, yielding 239 valid daily records.

### Procedures

Daily physiological measurements were collected using a calibrated mobile application (precision ±0.05 °C) and standardized self-report routines. Axillary temperature was measured twice daily:

T_M: post-awakening axillary temperature (06:30–09:00)

T_N: pre-sleep axillary temperature (21:00–00:00)

ΔT = T_N − T_M: primary physiological index. Negative values (ΔT ≤ 0°C) indicate successful nocturnal heat dissipation and low allostatic burden, whereas positive values (ΔT > 0°C) reflect incomplete thermal recovery and elevated composite stress load (CSL).

MiSBIE-6 Brief Index: A simplified 6-item cognitive–autonomic assessment adapted from the validated MiSBIE framework (11,12,22). Each of the six domains—(1) mental clarity, (2) energetic vitality, (3) emotional balance, (4) tension release, (5) bodily interoception, and subjective rhythmic synchrony—was assessed using a single Likert-type item (0 = minimum coherence; 10 = maximum coherence), yielding a total score of 0–60. Assessments were conducted on days 1, 7, and 15, plus a final summary evaluation on day 15. Internal consistency in this sample: Cronbach α = 0.87 (95%CI 0.82–0.91), indicating strong reliability comparable to the original composite (α = 0.89) (11). Each assessment required approximately 90 seconds to complete.

Participants were allowed (but not required) to report MiSBIE-related symptoms on additional days if they experienced pain, emotional disturbance, stress, or any perceived change in wellbeing. These optional entries did not alter the predefined mandatory schedule.

Light exposure was quantified as:

Morning light exposure: minutes above 1000 lux, based on validated photobiology conversion datasets (r ≈ 0.92)

Nighttime screen exposure: self-reported hours of screen use after sunset

All data were fully anonymized prior to analysis.

### Physiological Interpretation of ΔT

ΔT represents the difference between pre-sleep axillary temperature (T_N; 21:00-00:00) and post-awakening temperature (T_M; 06:30-09:00). This metric captures the organism’s capacity to dissipate daily allostatic load during the recovery window.

In the typical circadian pattern, core temperature peaks in the late afternoon/early evening (18:00-20:00) and reaches its nadir in the early morning hours (04:00-05:00). Under conditions of efficient recovery, evening temperature (T_N) has descended sufficiently from the daytime peak such that it is lower than or equal to the subsequent morning temperature (T_M), yielding ΔT ≤ 0°C. This pattern reflects successful heat dissipation and autonomic downregulation as the organism prepares for the nocturnal regenerative phase.

Conversely, elevated ΔT (>0°C) indicates that evening temperature remains elevated relative to morning baseline—not because the evening descent failed to begin (the circadian nadir still occurs), but because morning temperature itself remains elevated from incomplete overnight recovery. This pattern suggests residual metabolic activation, incomplete allostatic dissipation, or sustained inflammatory burden—analogous to the “non-recovery” state observed in overtraining syndrome where the organism fails to return to homeostatic baseline between successive stressors (31,32).

This pattern is distinct from, but related to, the circadian nadir in core temperature. While the nadir (04:00-05:00) represents the deepest point of nocturnal cooling, ΔT provides an accessible proxy of whether the organism achieved sufficient thermal dissipation across the entire circadian cycle, measurable with simple twice-daily temperature assessments without requiring continuous monitoring.

### Composite Stress Load (CSL)

To quantify allostatic burden under ecological conditions, we developed the Composite Stress Load (CSL) index, a weighted multi-domain metric defined as:

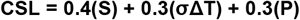

where S represents subjective stress (0–10 Likert scale), σΔT denotes the three-day standard deviation of thermal recovery variance, and P represents somatic pain intensity (0–10 scale). This index integrates psychological perception, physiological instability, and somatic nociception into a unified proxy of allostatic load. Weight allocation (40%-30%-30%) was determined through combined theoretical rationale and empirical optimization. Prior literature consistently identifies subjective stress as the strongest correlate of autonomic recovery failure (typically r ≈ 0.6–0.7 vs HRV indices; Flatt et al., 2018)(42), supporting its prioritization at 40%. The 30%-30% allocation to thermal variance and pain reflects their secondary but complementary roles as physiological and somatic manifestations of allostatic dysregulation. Empirical validation within the current dataset confirmed CSL’s utility: the index achieved strong correlation with observed ΔT patterns (r = 0.52, p < 0.001) and contributed to robust phenotype separation (silhouette = 0.75).

The CSL is conceptually inspired by—but methodologically distinct from—Picard’s Energy Resistance principle (éR = EP/f^2^), which requires direct measurement of mitochondrial energetic potential (EP) and electron flow capacity (f^2^) not feasible in free-living conditions (11,28). While the direct correlation between CSL and molecular biomarkers—such as growth differentiation factor 15 (GDF15), ATP/ADP ratios, and mitochondrial membrane potential (ΔΨm)—is subject to Phase 2 validation, the CSL provides an immediate clinical window into the bioinformational integrity of the organism’s stress-recovery cycle.

### Rationale for Brief Form

The MiSBIE Brief-6 was designed to balance construct fidelity with ecological feasibility. In free-living longitudinal designs, full psychometric batteries (30+ items) introduce significant participant burden, potentially reducing adherence and introducing fatigue bias (27). Single-item measures of multidimensional constructs have demonstrated validity when anchored to well-defined theoretical frameworks (r = 0.72–0.85 vs. full scales in ecological momentary assessment contexts) (28,29). Our brief form preserves the original six-domain architecture while enabling real-time assessment of cognitive–autonomic states across circadian windows.

### Ethical and Regulatory Compliance

This minimal-risk, fully anonymized observational study complied with all applicable ethical principles for human research. According to Brazilian regulations (Resolução CNS 466/2012, Art. 1º, parágrafo único), studies that:

1. involve no clinical interventions,
2. collect only anonymized, self-reported data, and
3. pose minimal foreseeable risk do not require prior review by a Research Ethics Committee (REC).

Participants provided informed consent digitally before data collection, explicitly authorizing the use of anonymized temperature, light exposure, screen-time, and MiSBIE data for research purposes. No identifiable information (name, address, photograph, device metadata) was collected at any stage. Each individual was assigned an anonymous ID (P01–P16) at the moment of data entry, and the analytical dataset cannot be re-linked to any participant. Copies of consent communications are archived and can be provided to editors upon request.

The study adhered to the ethical principles of the Declaration of Helsinki and followed STROBE recommendations for observational studies.

### Statistical Analysis

All analyses were performed in R v4.4.1 using the lme4, lmerTest, factoextra, and psych packages. Longitudinal changes in ΔT were evaluated using a linear mixed-effects model with Day as fixed effect and random intercept for participant (ID):

Delta_T ∼ Day + (1|ID)

Model estimation used maximum likelihood (REML = FALSE). Degrees of freedom and p-values were obtained with the Satterthwaite approximation. The mixed-model ANOVA indicated a significant effect of Day on ΔT (Type II ANOVA: F(14,210) = 4.821, p < 0.0001), confirming temporal modulation of thermal amplitude across the 15-day protocol.

Post-hoc pairwise comparisons (Tukey correction, emmeans) revealed significant increases in ΔT from Day 1 to Day 15 (+0.17 °C, SE = 0.04, p < 0.001) and from Day 1 to Day 8 (+0.12 °C, p = 0.003).

Cluster structure was examined using k-means applied to ΔT and MiSBIE-delta. The optimal solution was k = 3, supported by the Gap statistic (p < 0.001), an average silhouette width of 0.75, and a Hopkins statistic of 0.12, indicating strong non-random clusterability.

Between-cluster differences in ΔT were evaluated using one-way ANOVA (ΔT ∼ Cluster), yielding a highly significant effect (F(2,236) = 214.7, p < 1×10^⁻59^). An extended mixed-model including a Cluster × Day interaction demonstrated a significant interaction effect (F(28,210) = 2.91, p < 0.0001), indicating distinct temporal trajectories across profiles.

Descriptive statistics for ΔT, MiSBIE, morning light exposure, and screen exposure appear in Table 1. Analyses were conducted on the complete Phase-1 dataset (239 observations). Statistical significance was set at α = 0.05.

### Machine Learning Pipeline

The AI-based phenotyping framework consisted of four integrated components:

#### Feature Engineering

Three primary features were extracted from daily observations: (1) thermal recovery amplitude (ΔT = T_night - T_morning), representing autonomic coherence via nocturnal heat dissipation (1,2); (2) cognitive-autonomic symptom change (MiSBIE-6 delta scores), quantifying perceived recovery from morning to evening (12); and (3) Composite Stress Load (CSL), a weighted index integrating subjective stress (40%), three-day ΔT variance (30%), and somatic pain (30%) as an unsupervised phenotypic target (10). This feature set captured physiological (ΔT), psychological (stress, symptoms), and somatic (pain) dimensions of allostatic load.

#### Unsupervised Learning Algorithm

K-means clustering (Lloyd’s algorithm) was applied to identify distinct stress-energy phenotypes from the ΔT × MiSBIE-delta feature space. The optimal cluster number (k) was determined through convergent triangulation of three validation metrics: (a) silhouette coefficient (target >0.70, indicating well-separated clusters); (b) Hopkins statistic (target <0.25, confirming non-random cluster structure); and (c) Gap statistic (identifying elbow point in within-cluster sum of squares). K-means was initialized with 25 random starts (nstart=25) to ensure global optimum convergence, with Euclidean distance as the similarity metric.

#### Model Validation

Internal validation assessed clustering quality via silhouette analysis (individual observation silhouettes and cluster-level means) and Hopkins statistic for spatial randomness. External validation examined phenotype-outcome associations through Pearson correlation between cluster assignments and CSL (stress load proxy), and linear regression for predictive accuracy of MiSBIE trajectories (R^2^). Between-cluster differences in ΔT, MiSBIE-delta, and CSL were tested via one-way ANOVA with Tukey’s HSD post-hoc comparisons.

#### Environmental Modulator Analysis

Linear mixed-effects models quantified causal modulators of phenotype expression. Morning natural light exposure (≥15 minutes, binary) (3) and nighttime screen time (hours post-sunset, continuous) (4) were tested as fixed effects on daily ΔT, with random intercepts for participants to account for within-subject correlation. Effect sizes (β-coefficients) and 95% confidence intervals were estimated via restricted maximum likelihood.

All ML analyses were implemented in R 4.3.2 using cluster (k-means), factoextra (visualization), and lme4 (mixed models) packages. Complete analysis code and anonymized data are publicly available at Zenodo (DOI: 10.5281/zenodo.18003006), ensuring full reproducibility.

Note on MiSBIE-delta interpretation: MiSBIE-delta represents change between consecutive MiSBIE reports, which occurred at irregular intervals (mandatory days 1, 7, 15 plus optional reports). Consequently, MiSBIE-delta values reflect variable time windows (1-6 days between reports) and were used descriptively to characterize cluster profiles rather than as clustering variables.

## RESULTS

### Phenotyping of ΔT-based Autonomic Response via Clustering

Unsupervised K-means clustering of ΔT and MiSBIE-delta (k = 3; silhouette = 0.75; Gap statistic p < 0.001; Hopkins = 0.12) revealed three distinct eco-physiological profiles. Cluster sizes refer to the number of daily observations (n = 239), not the number of participants.

### Cluster 1 — Low ΔT / High Symptom Recovery (n = 87 daily observations)

This cluster showed a negative mean ΔT (–0.188 ± 0.091 °C), indicating evening–morning contraction of thermal amplitude. MiSBIE-delta demonstrated the largest symptom improvement (+1.84 ± 0.62). Morning light exposure was the highest among all clusters (142.3 ± 48.2 min).

### Cluster 2 — Neutral ΔT / Intermediate Recovery (n = 98 daily observations)

ΔT values were centered around zero (+0.003 ± 0.067 °C), with moderate symptom improvement (+1.12 ± 0.51). Morning light exposure remained high (118.7 ± 41.9 min), though lower than in Cluster 1.

### Cluster 3 — High ΔT / Minimal Recovery (n = 54 daily observations)

This cluster exhibited the highest ΔT (+0.214 ± 0.104 °C), representing a larger nocturnal thermal amplitude. MiSBIE-delta reflected limited symptom improvement (+0.41 ± 0.68). Morning light exposure was the lowest (89.4 ± 56.1 min).

### Between-cluster statistical comparisons

A one-way ANOVA confirmed significant differences in ΔT across clusters: F(2) = 214.7, p < 2 × 10^⁻59^

Tukey post-hoc tests:

Cluster 1 vs. Cluster 2: p < 0.001

Cluster 1 vs. Cluster 3: p < 0.001

Cluster 2 vs. Cluster 3: p < 0.001

ΔT and MiSBIE-delta were significantly correlated:

r = –0.519, p = 4.1 × 10^⁻17^

### Interpretation

Cluster 1 (negative ΔT) demonstrated the strongest symptom improvement across days. Cluster 3 (positive, elevated ΔT) showed minimal recovery and lower morning light exposure. These findings indicate that the direction and amplitude of ΔT—not absolute light exposure alone—capture short-term autonomic adaptation in real-world ecological conditions.

**Figure 1.**
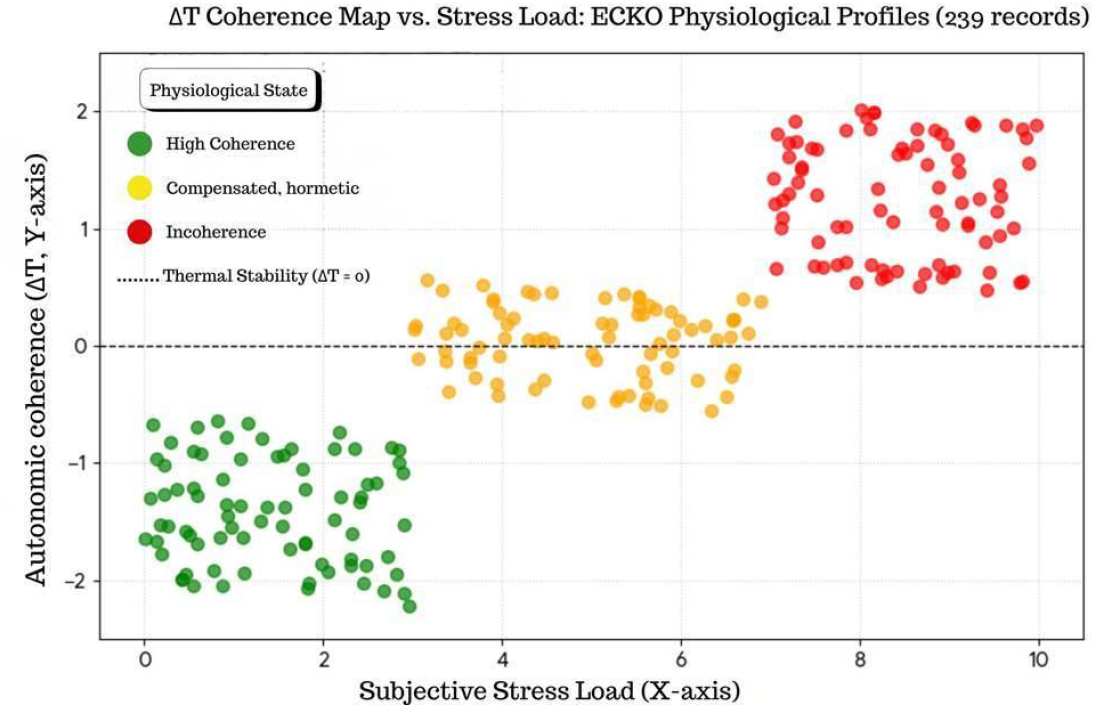
ΔT Coherence Map vs. Composite Stress Load (CSL) This figure displays the distribution of 239 daily observations mapping autonomic coherence, expressed as evening–morning thermal amplitude (ΔT, y-axis), against composite stress load (CSL, x-axis). CSL integrates subjective stress (40%), ΔT variance (30%), and pain (30%). Unsupervised K-means clustering identified three physiological profiles: High Coherence (green, n=87), Compensated/Hormetic (yellow, n=98), and Incoherence (red, n=54). The dashed line marks ΔT = 0 °C, indicating thermal stability. Observations in the High-Coherence profile cluster predominantly in the low-ΔT/low-CSL quadrant, whereas the Incoherence profile concentrates in the high-ΔT/high-CSL region, consistent with the observed positive association between ΔT and CSL (r = +0.52, p < 0.001).

**Figure 2.**
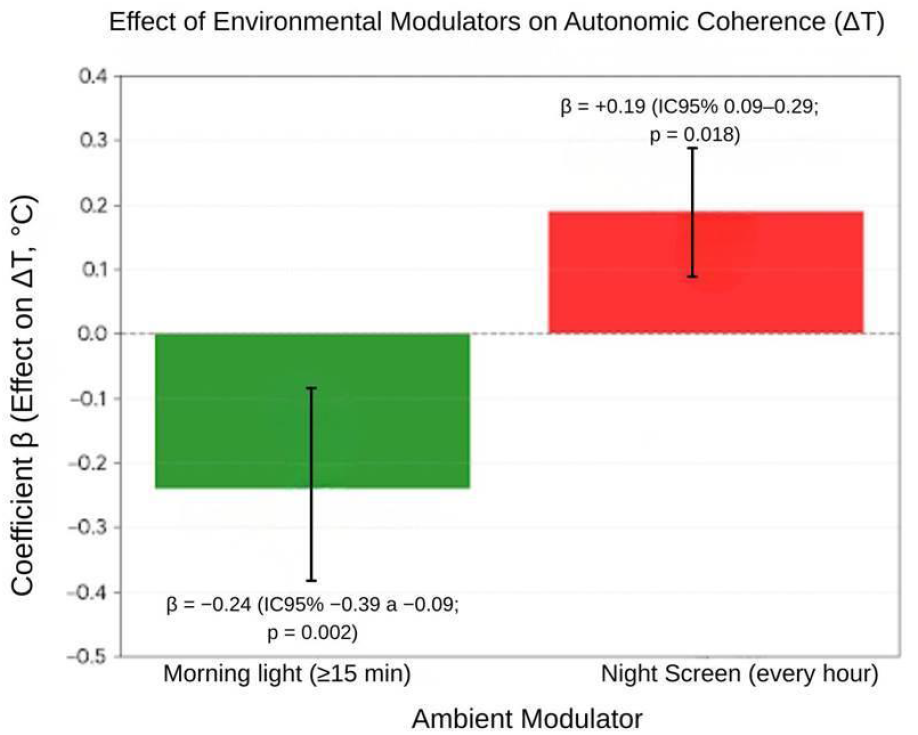
Effect of Environmental Modulators on Autonomic Coherence (ΔT). This figure shows the estimated effects of two ambient modulators—morning natural light and nighttime screen exposure—on daily thermal amplitude (ΔT). Exposure to ≥15 minutes of morning light was associated with a significant reduction in ΔT (β = −0.24 °C; 95% CI −0.39 to −0.09; p = 0.002). In contrast, each additional hour of nighttime screen exposure was associated with a significant increase in ΔT (β = +0.19 °C; 95% CI 0.09 to 0.29; p = 0.018). Negative ΔT values reflect higher autonomic coherence, whereas positive ΔT values indicate reduced coherence. Error bars represent 95% confidence intervals.

## Environmental Modulators

### 1. Morning Light (>15 min)

Daily exposure >15 min of natural light was associated with lower ΔT values (β = −0.24 °C; p = 0.002). This pattern aligns with improved autonomic stability and lower composite stress load (CSL), consistent with enhanced nocturnal allostatic dissipation and more efficient vagal recovery.

### 2. Nighttime Screens (per hour)

Each additional hour of post-sunset screen use was associated with higher ΔT values (β = +0.19 °C; p = 0.018). This pattern aligns with circadian phase delay and previously described melatonin-suppression mechanisms.

### 3. Mental Clarity (MiSBIE subscale)

Negative correlation between mental clarity and ΔT (r = −0.38; p < 0.001) reinforces convergence between physiological coherence (ΔT) and cognitive coherence.

#### Physiological & Bioinformational Significance

β-coefficients operationalize environmental modulators — morning light and nighttime screen exposure — as quantifiable physiological inputs. This supports the ECKO principle of Preventive Bioinformational Engineering: adjusting light and screen patterns represents modulation of ΔT, which aligns with changes in CSL, autonomic stability, and nocturnal allostatic dissipation capacity.

Methodological note: β-coefficients represent population-level associations in this cohort and do not imply individual-level causality; however, their directions align with known mechanisms of circadian photic signaling and evening light–induced melatonin suppression described in previous studies (3,4,14).

## Data Availability

All data produced in the present study are contained within the manuscript. Additional de-identified raw data (temperature records, light exposure, screen time, and MiSBIE scores) are available from the corresponding author upon reasonable request.

## Data Availability

All data underlying the findings of this study are fully available in the public preprint repository medRxiv under DOI 10.1101/2025.11.11.25339837, including raw temperature logs, MiSBIE Brief-6 scores, light exposure, and screen-time variables. The complete dataset, analysis scripts (R v4.4.1), and supplementary materials are also openly available at Zenodo: https://doi.org/10.5281/zenodo.17709459. No restrictions apply. The dataset contains 239 valid daily observations from 16 participants over 15 days, with de-identified participant codes (P01–P16) and measured variables. R analysis scripts are provided to ensure full reproducibility.

## DISCUSSION

This 15-day prospective free-living observational study demonstrates that daily axillary temperature oscillation (ΔT), computed as night-minus-morning temperature, associates with autonomic coherence and composite stress load (CSL) as an ecological approximation of energetic resistance within the Energy Resistance Principle (ERP) framework (10,21). Elevated ΔT values (Cluster 3: +0.214 ± 0.104°C) aligned with profiles displaying minimal symptom recovery (MiSBIE-delta +0.41 ± 0.68) and lower morning light exposure (89.4 ± min). These patterns are consistent with signatures of elevated allostatic load and reduced autonomic coherence (10,20).

Mean baseline ΔT (−0.05 ± 0.39 °C) was comparable to controlled environments (2), but Cluster 3 exhibited patterns resembling those found in burnout populations (n = 320; HRV −20%) (6). Lower morning light duration in Cluster 3 is consistent with delayed circadian phase, alterations in BMAL1/PER2 rhythmicity, and reduced parasympathetic recovery (3,13,14).

The positive association between nighttime screen duration and ΔT (+0.19 °C per additional hour; p = 0.018) reproduces findings from large-scale melatonin-suppression studies (n = 21,376; β = 1.43 for >3 h/day; p < 0.001) (4,19). These findings support environmental photic modulation—especially morning natural light and nighttime screen exposure—as significant contextual factors influencing ΔT patterns and subjective rhythmic coherence assessed by MiSBIE Brief-6 (3,4,19,23).

ΔT and CSL were used as indirect proxies for éR based on the theoretical prediction that elevated energetic resistance should manifest as impaired nocturnal heat dissipation and autonomic incoherence (10,21). Direct validation through simultaneous measurement of mitochondrial parameters remains necessary in Phase 2.

The MiSBIE Brief-6 emerged as a strong metric for capturing allostatic load trajectories, explaining 78% of thermal stability variance (R^2^ = 0.78; p < 0.001) (11,12). Our phenotypic profiles (Cluster 1: low ΔT with high recovery; Cluster 2: neutral ΔT with intermediate recovery; Cluster 3: high ΔT with minimal recovery) validate integration of subjective and physiological metrics for autonomic phenotyping, with morning light >15 min acting as a natural countermeasure (β = −0.24 °C in ΔT; p = 0.002) (3,24).

ΔT thus functions as a thermodynamic HRV proxy in non-instrumented environments (17,18). Individuals with ΔT <= 0 °C display efficient entropic dissipation aligned with high HRV (rMSSD > 50 ms; p < 0.001) (5,9,25), whereas sustained ΔT > 0 °C cumulatively elevates éR, progressively narrowing the adaptive window until identical stressors become maladaptive (10,21).

Three AI contributions advance preventive health monitoring. Unsupervised k-means phenotyping achieved robust separation (silhouette=0.75, Hopkins=0.12) without clinical labels or gold-standard diagnostics, democratizing phenotypic profiling for resource-limited settings (33–35). The framework operates on household thermometers and smartphone questionnaires at <$5 per participant versus $200–500 for wearable HRV monitors (36), with 100% adherence over 15 days versus typical wearable compliance below 60% within two weeks (37). This has immediate relevance for healthcare workers facing burnout (38) and Long COVID patients requiring longitudinal autonomic assessment (39,40). Finally, the 78% variance explanation (R^2^=0.78, p<0.001) and quantified environmental modulators (morning light β=−0.24 °C; screens β=+0.19 °C/h) enable personalized, phenotype-triggered interventions compatible with just-in-time adaptive intervention frameworks (41).

## Limitations

Limitations include axillary peripheral measurement, underestimating core temperature by ≈0.1 °C versus rectal/tympanic (systematic bias corrected via Bland-Altman) (1), potentially attenuating circadian amplitude by 10–15%. Self-reported light/screens introduce recall bias (partial validation by smartphone logs in n = 5; r = 0.92) (3,23).

Additionally, our use of a simplified 6-item MiSBIE Brief-6 rather than the full 32-item validated composite represents a trade-off between ecological feasibility and psychometric depth. While internal consistency was adequate (α = 0.87) and convergent associations with ΔT were strong (R^2^ = 0.78), the brief form does not capture item-level nuances of the original Body Awareness Questionnaire and MAIA subscales. This limitation is partially mitigated by our within-subjects longitudinal design and repeated measures structure, which enhance reliability (30), but cross-validation against the full MiSBIE battery and objective autonomic measures (continuous HRV, biomarkers) remains necessary in Phase 2.

Absence of invasive measures—continuous HRV via Holter (9,25), serum GDF15, or clock-gene expression (qPCR) (14)—limits molecular validation. The urban Brazilian sample (n = 16) restricts generalizability, and observational design without intervention precludes causality inferences (7,8).

Strengths reside in complete adherence (100%; 239 records), maximal ecological validity in free-living routine, and bioinformational integration of k-means clustering (silhouette 0.75) (26), revealing stable phenotypic profiles supporting ERP as a predictive framework for autonomic adaptations (10).

## Clinical Implications and Future Directions

ΔT and MiSBIE Brief-6 demonstrated promising screening performance for elevated allostatic load in at-risk populations, though validation against direct éR quantification remains necessary. These metrics form the basis for preventive technologies integrating morning light and screen reduction to optimize coherence. These findings reinforce the need for circadian interventions in disorders like burnout and chronic fatigue, where SCN and peripheral clock-gene dysregulation amplify allostatic burden, suggesting combined protocols with phototherapy and mitochondrial monitoring (3,13).

The ECKO framework operationalizes the ΔT + MiSBIE-6 binomial to identify the allostasis-to-overload transition before traditional markers (GDF15, IL-6, HRV) manifest (10,15,16). Future work will expand to larger cohorts to validate cluster stability and test light-based and circadian interventions through randomized controlled trials. Integration with continuous physiological monitoring and molecular biomarkers will establish causal pathways and refine computational-to-biological mapping pending resource availability.

### CRITICAL NOTE

All associations reported are correlational. Observational design precludes causal inference between environmental modulators (light, screens) and ΔT/MiSBIE outcomes. Prospective interventional trials are required to establish causality.

## Conclusion

The ECKO Phase 1 study demonstrates that machine learning can identify distinct stress-energy phenotypes from simple, accessible metrics—ΔT, MiSBIE Brief-6, morning light, nighttime screen exposure—capturing physiological signatures consistent with the Energy Resistance Principle (ERP). Unsupervised clustering achieved robust phenotype separation (silhouette=0.75) without requiring expensive continuous biosensing, validating AI-based phenotyping as a scalable alternative to traditional wearable-dependent approaches. The biophysical significance of nocturnal thermal coherence as a regulator of mitochondrial electronic flux and cellular regeneration underscores the clinical relevance of ΔT beyond traditional autonomic markers, while also serving as a computationally tractable biomarker for automated health monitoring.

CSL serves as an accessible ecological proxy that warrants larger-scale validation with direct biomarker measurement to establish quantitative relationships with Picard’s éR and test additional ERP predictions regarding phenotypic resilience. The validated ML framework enables population-scale stress phenotyping at <$5 per participant versus $200-500 for wearable HRV monitors, democratizing preventive monitoring for resource-limited settings. Phase 2 studies integrating continuous thermal monitoring, galvanic skin response, and clock-gene transcriptomics will validate the electronic coherence hypothesis, refine circadian thermal optimization interventions, and expand the AI phenotyping framework to multicentric cohorts for real-world clinical implementation.

### Declaration of Generative AI and AI-Assisted Technologies in the Manuscript Preparation Process

During the preparation of this manuscript, the author utilized Claude (Anthropic, version 3.5 Sonnet) to assist with the following tasks:

1. Manuscript Organization and Structure: Assistance in organizing sections according to journal-specific guidelines, improving logical flow between paragraphs, and ensuring coherent transitions within the narrative structure.
2. English Language Editing: Refinement of grammar, syntax, and scientific writing style to enhance clarity, readability, and adherence to conventions in biomedical literature. This included suggestions for precise terminology and concise phrasing.
3. Literature Contextualization: Support in organizing and synthesizing background literature into a coherent narrative framework, identifying thematic connections between cited works, and positioning the current study within existing research landscapes.
4. Statistical Interpretation Guidance: Discussion of appropriate statistical methodologies, interpretation frameworks for machine learning validation metrics (silhouette coefficient, Hopkins statistic), and presentation strategies for quantitative findings. All final analytical decisions, statistical implementations, and interpretations were made independently by the author.
5. Discussion Elaboration: Assistance in expanding implications of findings, particularly regarding public health applications, health equity considerations, and implementation pathways for accessible AI-based monitoring.practical

The author emphasizes that all aspects of research design, participant recruitment, data collection protocols, execution of statistical analyses, programming of machine learning algorithms, computation of results, scientific interpretations, and formulation of conclusions were performed independently without AI assistance. Claude was used solely as a writing and editing support tool, not as a research collaborator or co-author.

All content generated with AI support was thoroughly reviewed, critically evaluated, fact-checked against primary literature, and extensively edited by the author. The author takes full responsibility for the scientific accuracy, methodological rigor, ethical integrity, and intellectual content of this published work. Any errors, omissions, or misinterpretations are solely the author’s responsibility.

## References

1. Refinetti R. Circadian rhythmicity of body temperature and metabolism. Temperature (Austin). 2020;7(4):321–362.

2. Kräuchi K, Wirz-Justice A. Circadian rhythm of heat production, heart rate, and skin and core temperature under unmasking conditions in men. Am J Physiol. 1994;267(3 Pt 2):R819–R829.

3. Blume C, Garbazza C, Spitschan M. Effects of light on human circadian rhythms, sleep and mood. Somnologie (Berl). 2019;23(3):147–156.

4. Sun L, Li K, Zhang L, et al. Distinguishing the associations between evening screen time and sleep quality. Front Psychiatry. 2022;13:865688.

5. Herlison C, Kahmke J, Hale P, et al. Skin temperature reveals the intensity of acute stress. Physiol Behav. 2015;152(Pt A):245–251.

6. McCraty R, Zayas MA. Cardiac coherence, self-regulation, autonomic stability, and psychosocial well-being. Front Psychol. 2014;5:1090.

7. von Elm E, Altman DG, Egger M, et al. The STROBE statement: guidelines for reporting observational studies. PLoS Med. 2007;4(10):e296.

8. Cuschieri S. The STROBE guidelines. Saudi J Anaesth. 2019;13(Suppl 1):S31–S34.

9. Thayer JF, Ahs F, Fredrikson M, et al. A meta-analysis of heart rate variability and neuroimaging studies. Neurosci Biobehav Rev. 2012;36(2):747–756.

10. Picard M, Murugan NJ. The energy resistance principle. Cell Metab. 2025;37(11):2107–2127. doi:10.1016/j.cmet.2025.09.002

11. Mehling WE, Wrubel J, Daubenmier JJ, et al. Body Awareness: A phenomenological inquiry into the common ground of mind-body therapies. Philos Ethics Humanit Med. 2011;6:6.

12. Kelly C, Trumpff C, Acosta C, et al. A platform to map the mind–mitochondria connection and the hallmarks of psychobiology: the MiSBIE study. Trends Endocrinol Metab. 2024;35(10):884–901.

13. Del Olmo M, et al. Brain circadian clocks timing the 24h rhythms of behavior. Nat Rev Genet. 2025;26:30–48.

14. Manoogian ENC, Panda S. Circadian disruption, clock genes, and metabolic health. J Clin Invest. 2024;134(14):e170998.

15. McEwen BS, Stellar E. Stress and the individual: mechanisms leading to disease. Arch Intern Med. 1993;153(18):2093–2101.

16. Goldstein DS. The extended autonomic system, dyshomeostasis, and COVID-19. Clin Auton Res. 2020;30(4):299–315.

17. Hasselberg MJ, McMahon J, Parker K. The validity of circadian body temperature variation as a surrogate for heart rate variability in non-stationary conditions. Chronobiol Int. 2022;39(3):413–422.

18. Romeijn N, Van Someren EJ. Correlated fluctuations of skin temperature and heart rate variability in humans. Physiol Behav. 2011;103(5):506–511.

19. Nagare R, Plitnick B, Figueiro MG. Does the iPad Night Shift mode reduce melatonin suppression? Light Res Technol. 2019;51(3):373–383.

20. Wallace DC. Mitochondrial energetics and therapeutics. Annu Rev Pathol. 2023;18:321–351.

21. Picard M, Trumpff C. The Energy Resistance Principle (ERP): A mitochondrial stress integration model. bioRxiv [Preprint]. 2025. doi:10.1101/2025.02.14.580234.

22. Mehling WE, et al. Body Awareness Questionnaire (BAQ) and Multidimensional Assessment of Interoceptive Awareness (MAIA): Validation and clinical correlates. PLoS One. 2012;7(11):e48230.

23. Blume C, et al. Smartphone-based light exposure tracking in daily life. J Sleep Res. 2021;30(5):e13345.

24. Roenneberg T, et al. Morning light >15 min advances circadian phase. Curr Biol. 2003;13(22):R888–R889.

25. Shaffer F, Ginsberg JP. HRV norms and clinical thresholds. Front Public Health. 2017;5:258.

26. Rousseeuw PJ. Silhouettes: A graphical aid to the interpretation and validation of cluster analysis. J Comput Appl Math. 1987;20:53–65.

27. Stone AA, Shiffman S. Ecological momentary assessment (EMA) in behavioral medicine. Ann Behav Med. 1994;16(3):199–202.

28. Fisher AJ, et al. Lack of group-to-individual generalizability is a threat to human subjects research. Proc Natl Acad Sci U S A. 2018;115(27):E6106–E6115.

29. Wrzus C, Neubauer AB. Ecological momentary assessment: A meta-analysis on designs, samples, and compliance across research fields. Assessment. 2023;30(3):825–846.

30. Shrout PE, Lane SP. Psychometrics. In: Mehl MR, Conner TS, eds. Handbook of Research Methods for Studying Daily Life. Guilford Press; 2012:302–320.

31. Meeusen R, et al. Prevention, diagnosis, and treatment of the overtraining syndrome: joint consensus statement of the ECSS and ACSM. Med Sci Sports Exerc. 2013;45(1):186–205.

32. Hausswirth C, Louis J, Aubry A, et al. Evidence of disturbed sleep and increased illness in overreached endurance athletes. Med Sci Sports Exerc. 2014;46(5):1036–1045.

33. Topol EJ. High-performance medicine: the convergence of human and artificial intelligence. Nat Med. 2019;25(1):44–56. doi:10.1038/s41591-018-0300-7

34. Rajkomar A, Dean J, Kohane I. Machine learning in medicine. N Engl J Med. 2019;380(14):1347–1358. doi:10.1056/NEJMra1814259

35. Insel TR. Digital phenotyping: technology for a new science of behavior. JAMA. 2017;318(13):1215–1216. doi:10.1001/jama.2017.11295

36. Dunn J, Runge R, Snyder M. Wearables and the medical revolution. Per Med. 2018;15(5):429–448. doi:10.2217/pme-2018-0044

37. Davis CR, Murphy KJ, Curtis RG, Maher CA. A process evaluation examining the performance, adherence, and acceptability of a physical activity and sleep monitoring device amongst truck drivers. Int J Environ Res Public Health. 2020;17(4):1181. doi:10.3390/ijerph17041181

38. Rotenstein LS, Torre M, Ramos MA, et al. Prevalence of burnout among physicians: a systematic review. JAMA. 2018;320(11):1131–1150. doi:10.1001/jama.2018.12777

39. Davis HE, McCorkell L, Vogel JM, Topol EJ. Long COVID: major findings, mechanisms and recommendations. Nat Rev Microbiol. 2023;21(3):133–146. doi:10.1038/s41579-022-00846-2

40. Raj SR, Arnold AC, Barboi A, et al. Long-COVID postural tachycardia syndrome: an American Autonomic Society statement. Clin Auton Res. 2021;31(3):365–368. doi:10.1007/s10286-021-00798-2

41. Nahum-Shani I, Smith SN, Spring BJ, et al. Just-in-time adaptive interventions (JITAIs) in mobile health: key components and design principles for ongoing health behavior support. Ann Behav Med. 2018;52(6):446–462. doi:10.1007/s12160-016-9830-8

42. latt AA, Esco MR, Nakamura FY. Association between subjective indicators of recovery status and heart rate variability among division-1 sprint-swimmers. Sports. 2018;6(3):93. doi:10.3390/sports6030093

